# Genetic mechanisms of critical illness in Covid-19

**DOI:** 10.1101/2020.09.24.20200048

**Authors:** Erola Pairo-Castineira, Sara Clohisey, Lucija Klaric, Andrew Bretherick, Konrad Rawlik, Nick Parkinson, Dorota Pasko, Susan Walker, Anne Richmond, Max Head Fourman, Clark D Russell, Andrew Law, James Furniss, Elvina Gountouna, Nicola Wrobel, Loukas Moutsianas, Bo Wang, Alison Meynert, Zhijian Yang, Ranran Zhai, Chenqing Zheng, Fiona Griffiths, Wilna Oosthuyzen, Graeme Grimes, Barbara Shih, Sean Keating, Marie Zechner, Chris Haley, David J. Porteous, Caroline Hayward, Julian Knight, Charlotte Summers, Manu Shankar-Hari, Paul Klenerman, Lance Turtle, Antonia Ho, Charles Hinds, Peter Horby, Alistair Nichol, David Maslove, Lowell Ling, Danny McAuley, Hugh Montgomery, Timothy Walsh, The GenOMICC Investigators, The ISARIC-4C Investigators, The Covid-19 Human Genetics Initiative, Xia Shen, Kathy Rowan, Angie Fawkes, Lee Murphy, Chris P. Ponting, Albert Tenesa, Mark Caulfield, Richard Scott, Peter J.M. Openshaw, Malcolm G. Semple, Veronique Vitart, James F. Wilson, J. Kenneth Baillie

**Affiliations:** Roslin Institute, University of Edinburgh, Easter Bush, Edinburgh, EH25 9RG, UK; MRC Human Genetics Unit, Institute of Genetics and Molecular Medicine, University of Edinburgh, Western General Hospital, Crewe Road, Edinburgh, EH4 2XU, UK; Genomics England, London, UK; University of Edinburgh Centre for Inflammation Research, The Queen’s Medical Research Institute, Edinburgh, UK; Centre for Genomic and Experimental Medicine, Institute of Genetics and Molecular Medicine, University of Edinburgh, Western General Hospital, Crewe Road, Edinburgh, EH4 2XU, UK; Edinburgh Clinical Research Facility, Western General Hospital, University of Edinburgh, EH4 2XU, UK; Biostatistics Group, School of Life Sciences, Sun Yat-sen University, Guangzhou, China; Intenstive Care Unit, Royal Infirmary of Edinburgh, 54 Little France Drive, Edinburgh, EH16 5SA, UK; Wellcome Centre for Human Genetics, University of Oxford, Oxford, UK; Department of Medicine, University of Cambridge, Cambridge, UK; Department of Intensive Care Medicine, Guy’s and St. Thomas NHS Foundation Trust, London, UK; School of Immunology and Microbial Sciences, King’s College London, UK; NIHR Health Protection Research Unit for Emerging and Zoonotic Infections, Institute of Infection, Veterinary and Ecological Sciences University of Liverpool, Liverpool, L69 7BE, UK; MRC-University of Glasgow Centre for Virus Research, Institute of Infection, Immunity and Inflammation, College of Medical, Veterinary and Life Sciences, University of Glasgow, Glasgow, UK; William Harvey Research Institute, Barts and the London School of Medicine and Dentistry, Queen Mary University of London, London EC1M 6BQ, UK; Centre for Tropical Medicine and Global Health, Nuffield Department of Medicine, University of Oxford, Old Road Campus, Roosevelt Drive, Oxford, OX3 7FZ, UK; Clinical Research Centre at St Vincent’s University Hospital, University College Dublin, Dublin, Ireland; Australian and New Zealand Intensive Care Research Centre, Monash University, Melbourne, Australia; Intensive Care Unit, Alfred Hospital, Melbourne, Australia; Department of Critical Care Medicine, Queen’s University and Kingston Health Sciences Centre, Kingston, ON, Canada; Department of Anaesthesia and Intensive Care, The Chinese University of Hong Kong, Prince of Wales Hospital, Hong Kong, China; Wellcome-Wolfson Institute for Experimental Medicine, Queen’s University Belfast, Belfast, Northern Ireland, UK; Department of Intensive Care Medicine, Royal Victoria Hospital, Belfast, Northern Ireland, UK; UCL Centre for Human Health and Performance, London, W1T 7HA, UK; Centre for Global Health Research, Usher Institute of Population Health Sciences and Informatics, Teviot Place, Edinburgh EH8 9AG, UK; Department of Medical Epidemiology and Biostatistics, Karolinska Institutet, Stockholm, Sweden; Intensive Care National Audit & Research Centre, London, UK; Great Ormond Street Hospital for Children NHS Foundation Trust, London, UK; National Heart & Lung Institute, Imperial College London (St Mary’s Campus), Norfolk Place, Paddington, London W2 1PG, UK; University of Liverpool, Liverpool, UK

## Abstract

The subset of patients who develop critical illness in Covid-19 have extensive inflammation affecting the lungs^1^ and are strikingly different from other patients: immunosuppressive therapy benefits critically-ill patients, but may harm some non-critical cases.^2^ Since susceptibility to life-threatening infections and immune-mediated diseases are both strongly heritable traits, we reasoned that host genetic variation may identify mechanistic targets for therapeutic development in Covid-19.^3^

GenOMICC (Genetics Of Mortality In Critical Care, genomicc.org) is a global collaborative study to understand the genetic basis of critical illness. Here we report the results of a genome-wide association study (GWAS) in 2244 critically-ill Covid-19 patients from 208 UK intensive care units (ICUs), representing >95% of all ICU beds. Ancestry-matched controls were drawn from the UK Biobank population study and results were confirmed in GWAS comparisons with two other population control groups: the 100,000 genomes project and Generation Scotland.

We identify and replicate three novel genome-wide significant associations, at chr19p13.3 (rs2109069, *p* = 3.98 × 10^−12^), within the gene encoding dipeptidyl peptidase 9 (*DPP9*), at chr12q24.13 (rs10735079, *p* =1.65 × 10^−8^) in a gene cluster encoding antiviral restriction enzyme activators (*OAS1, OAS2, OAS3*), and at chr21q22.1 (rs2236757, *p* = 4.99 × 10^−8^) in the interferon receptor gene *IFNAR2*. Consistent with our focus on extreme disease in younger patients with less comorbidity, we detect a stronger signal at the known 3p21.31 locus than previous studies (rs73064425, *p* = 4.77 × 10^−30^).

We identify potential targets for repurposing of licensed medications. Using Mendelian randomisation we found evidence in support of a causal link from low expression of *IFNAR2*, and high expression of *TYK2*, to life-threatening disease. Transcriptome-wide association in lung tissue revealed that high expression of the monocyte/macrophage chemotactic receptor *CCR2* is associated with severe Covid-19.

Our results identify robust genetic signals relating to key host antiviral defence mechanisms, and mediators of inflammatory organ damage in Covid-19. Both mechanisms may be amenable to targeted treatment with existing drugs. Large-scale randomised clinical trials will be essential before any change to clinical practice.

## Introduction

Critical illness in Covid-19 is caused, in part, by inflammatory injury affecting the lungs and lung blood vessels.^4,5^ There are therefore at least two distinct biological components to mortality risk: susceptibility to viral infection, and propensity to develop harmful pulmonary inflammation. Susceptibility to life-threatening infections^6^ and immune-mediated diseases are both strongly heritable. In particular, susceptibility to respiratory viruses^7^ such as influenza^8^ is heritable and known to be associated with specific genetic variants.^9^ In Covid-19, one genetic locus, 3p21.31 has been repeatedly associated with hospitalisation.^10,11^ As with other viral illnesses,^12^ there are several examples of loss-of-function variants affecting essential immune processes that lead to severe disease in young people: for example *TLR7* defects among 4 cases with severe disease.^13^ Understanding the molecular mechanisms of critical illness in Covid-19 may reveal new therapeutic targets to modulate this host immune response to promote survival.^3^

There is now strong evidence that critical illness caused by Covid-19 is qualitatively different from mild or moderate disease, even among hospitalised patients. There are multiple distinct disease phenotypes with differing patterns of presenting symptoms^14^ and marked differential responses to immunosuppressive therapy.^2^ In patients without respiratory failure, there is a trend towards harm from treatment with corticosteroids, whereas among patients with critical respiratory failure, there is a very substantial benefit.^2^ On this basis, we can consider patients with critical Covid-19 respiratory failure to have distinct pathophysiology.

In the UK, the group of patients admitted to critical care is relatively homogeneous, with profound hypoxaemic respiratory failure being the archetypal presentation.^15^ The active disease process in these patients is strikingly responsive to corticosteroid therapy^16^ and is characterised by pulmonary inflammation including diffuse alveolar damage, macrophage/monocyte influx, mononuclear cell pulmonary artery vasculitis and microthrombus formation.^4,5^

Host-directed therapies have long been an aspiration for the treatment of severe disease caused by respiratory viruses.^17^ Identification of genetic loci associated with susceptibility to Covid-19 may lead to specific targets for repurposing or drug development.^3^

The GenOMICC (Genetics Of Mortality In Critical Care, genomicc.org) study has been recruiting patients with critical illness syndromes, including influenza, sepsis, and emerging infections, for 5 years. GenOMICC works in partnership with Genomics England to study Covid-19. In order to better understand the host mechanisms leading to life-threatening Covid-19, we performed a genome-wide association study comparing to controls from population genetic studies in the UK.

## Results

Critically-ill cases were recruited through the GenOMICC study in 208 UK Intensive Care Units and hospitalised cases through the International Severe Acute Respiratory Infection Consortium (ISARIC) Coronavirus Clinical Characterisation Consortium (4C) study. Demographic and summary clinical characteristics of the cohort are described in Table 1. Cases were representative of the UK critically-ill population.^15^

**Table 1:**
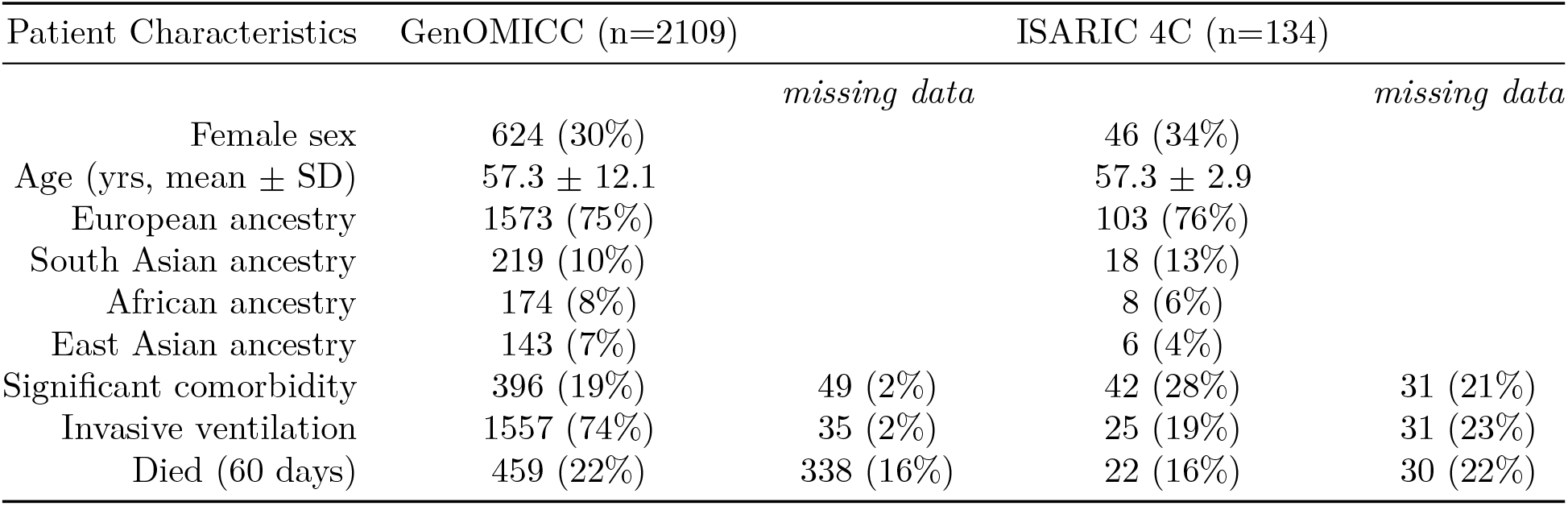
Baseline characteristics of patients included. Significant comorbidity was defined as the presence of functionally-limiting comorbid illness in GenOMICC, in the assessment of the treating clinicians. In ISARIC 4C significant comorbidity refers to the presence of any chronic cardiac, lung, kidney, or liver disease, cancer or dementia. Age is shown as mean ± standard deviation.

DNA was extracted from whole blood and genome-wide genotyping and quality control were performed according to standard protocols (Materials & Methods). Briefly, genetic ancestry was inferred for unrelated individuals passing quality control using ADMIXTURE and reference individuals from the 1000 Genomes project. Imputation was performed using the TOPMed reference panel.^18^ Whole genome sequencing was performed on a subset of 1613 cases, and used to confirm both array-genotyped and imputed genotypes. From the 4469187 imputed variants that passed all filters after GWAS, 72658 did not pass QC filtering in WGS data and were removed. Comparing the allele frequencies of each SNP between WGS and imputation, the correlation of allele frequencies was r^2^=0.9994. All variants with a difference of > 5% were removed from the analysis, leaving 4396207 imputed variants.

Ancestry-matched controls not having Covid-19 PCR tests were selected from the large population-based cohort UK Biobank in a ratio of 5 controls to 1 case. GWAS was carried out separately by ancestry group using logistic regression in PLINK and accounting for age, sex, postal code deprivation decile and principal components of ancestry. As well as standard filters for minor allele frequency (>0.01), imputation quality (0.9) and Hardy-Weinberg equilibrium (10^−^50^), GWAS results were filtered on allele frequency against the genome aggregation database (gnomAD), to avoid biases arising from different imputation panels (and arrays) between cases and controls. The largest ancestry group contained 1676 individuals of European descent (EUR).

Following linkage disequilibrium-based clumping, 15 independent association signals were genome-wide significant at p < 5 × 10^−8^. Eight of these were successfully validated using a GWAS using two independent population genetic studies (100,000 genomes and Generation Scotland) as controls (Table 2).

**Table 2:** Lead variants from independent genome-wide significant regions. Full summary statistics are provided in Supplementary Information. chr:pos - chromosome and position of the top SNP (build 37); Risk – risk allele; Other - other allele; RAF - risk allele frequency; OR - effect size (odds ratio) of the risk allele; locus – gene nearest to the top SNP.

| SNP | chr:pos(b37) | Risk | Other | RAF | OR | P | Locus |
| --- | --- | --- | --- | --- | --- | --- | --- |
| rs73064425 | 3:45901089 | T | C | 0.08 | 2.14 | $4.77 \times 10^{-30}$ | *LZTFL1* |
| rs9380142 | 6:29798794 | A | G | 0.70 | 1.30 | $3.23 \times 10^{-8}$ | *HLA-G* |
| rs143334143 | 6:31121426 | A | G | 0.079 | 1.85 | $8.82 \times 10^{-18}$ | *CCHCR1* |
| rs3131294 | 6:32180146 | G | A | 0.87 | 1.45 | $2.81 \times 10^{-8}$ | *NOTCH4* |
| rs10735079 | 12:113380008 | A | G | 0.64 | 1.29 | $1.65 \times 10^{-8}$ | *OAS3* |
| rs74956615 | 19:10427721 | A | T | 0.06 | 1.59 | $2.31 \times 10^{-8}$ | *ICAM5/TYK2* |
| rs2109069 | 19:4719443 | A | G | 0.33 | 1.36 | $3.98 \times 10^{-12}$ | *DPP9* |
| rs2236757 | 21:34624917 | A | G | 0.29 | 1.28 | $5.00 \times 10^{-8}$ | *IFNAR2* |

### GWAS results

Since no study of critical illness in Covid-19 of sufficient size is available, replication was sought in the Covid-19 Host Genetics Initiative (HGI) hospitalised COVID-19 versus population analysis, with UK Biobank cases excluded. In addition to the locus on chr3 already reported (rs73064425, OR=2.14, discovery p=4.77 × 10^−30^), we found robust replication for the novel associations in three loci from GenOMICC: a locus on chr12 in the *OAS* gene cluster (rs74956615, OR=1.59, discovery p= 1.65 × 10^−8^) and in *DPP9* on chr19 (rs2109069, OR=1.36, discovery p=3.98 × 10^−12^) and locus on chromosome 21, containing gene IFNAR2 (rs2236757, OR=1.28, p=5 × 10^−8^) (Table 3).

**Table 3:** Replication in external data from Covid-19 HGI. chr:pos - chromosome and position of the top SNP (build 37); Risk – risk allele; Other - other allele; beta - effect size of the risk allele; OR - odds ratio; locus – gene nearest to the top SNP; gcc.eur - GenOMICC study, European ancestry; hgi - Covid-19 human genetics initiative; ^*^ Bonferroni significant.

| SNP | chr:pos (b37) | Risk | Other | beta gcc.eur | P gcc.eur | beta hgi | P hgi | Locus |
| --- | --- | --- | --- | --- | --- | --- | --- | --- |
| rs73064425 | 3:45901089 | T | C | 0.76 | $4.8 \times 10^{-30}$ | 0.53 | $1.6 \times 10^{-12*}$ | *LZTFL1* |
| rs9380142 | 6:29798794 | A | G | 0.26 | $3.2 \times 10^{-8}$ | 0.036 | 0.46 | *HLA-G* |
| rs143334143 | 6:31121426 | A | G | 0.62 | $8.8 \times 10^{-18}$ | 0.064 | 0.34 | *CCHCR1* |
| rs3131294 | 6:32180146 | G | A | 0.38 | $2.8 \times 10^{-8}$ | 0.1 | 0.2 | *NOTCH4* |
| rs10735079 | 12:113380008 | A | G | 0.26 | $1.6 \times 10^{-8}$ | 0.14 | 0.0027* | *OAS3* |
| rs2109069 | 19:4719443 | A | G | 0.31 | $4 \times 10^{-12}$ | 0.17 | 0.00031* | *DPP9* |
| rs74956615 | 19:10427721 | A | T | 0.46 | $2.3 \times 10^{-8}$ | 0.14 | 0.4 | *ICAM5/TYK2* |
| rs2236757 | 21:34624917 | A | G | 0.25 | $5 \times 10^{-8}$ | 0.18 | 0.00016* | *IFNAR2* |

### Replication

To further increase power for exploratory downstream analyses meta-analysis of GENOMICC and HGI was performed using inverse-variance meta-analysis in METAL,^20^ discovering 2 additional loci, one on chromosome 8, near *HAS2* (rs10087754, OR=1.19, meta-analysis p = 3.1 × 10^−8^) and another on chromosome 19, near *TYK2* (rs11085727, OR=1.25, p = 1.57 × 10^−10^) (Table 4).

**Table 4:** Meta-analysis of GenOMICC (EUR) and HGI hospitalized covid vs. population studies. SNP – the strongest SNP in the locus, ; A1 – effect allele; A2 - alternative allele; beta - effect size of the effect allele; nearest gene – gene nearest to the top SNP.

| SNP | A1 | A2 | beta eur | p eur | beta hgi.meta | OR hgi.meta | p hgi.meta | Gene |
| --- | --- | --- | --- | --- | --- | --- | --- | --- |
| rs67959919 | A | G | 0.76 | $7.1 \times 10^{-30}$ | 0.66 | 1.9 | $7.3 \times 10^{-38}$ | *LZTFL1* |
| rs143334143 | A | G | 0.62 | $8.8 \times 10^{-18}$ | 0.31 | 1.4 | $7 \times 10^{-10}$ | *CCHCR1* |
| rs9501257 | G | A | 0.49 | $1.1 \times 10^{-11}$ | 0.31 | 1.4 | $3.8 \times 10^{-9}$ | *HLA-DPB1* |
| rs622568 | C | A | 0.45 | $7.1 \times 10^{-16}$ | 0.29 | 1.3 | $7 \times 10^{-11}$ | *VSTM2A* |
| rs10087754 | T | A | 0.19 | $7.6 \times 10^{-6}$ | 0.18 | 1.2 | $3.1 \times 10^{-8}$ | *HAS2-ASI* |
| rs10860891 | C | A | 0.55 | $1.6 \times 10^{-18}$ | 0.3 | 1.3 | $7.5 \times 10^{-10}$ | *IGF1* |
| rs4766664 | G | T | 0.25 | $3.1 \times 10^{-8}$ | 0.2 | 1.2 | $2.5 \times 10^{-9}$ | *OAS1* |
| rs2277732 | A | C | 0.3 | $1.8 \times 10^{-11}$ | 0.25 | 1.3 | $1.6 \times 10^{-13}$ | *DPP9* |
| rs11085727 | T | C | 0.24 | $1.3 \times 10^{-7}$ | 0.22 | 1.2 | $1.6 \times 10^{-10}$ | *TYK2* |
| rs13050728 | T | C | 0.23 | $3 \times 10^{-7}$ | 0.21 | 1.2 | $2.5 \times 10^{-10}$ | *IFNAR2* |

#### Mendelian randomisation

Mendelian randomisation provides evidence for a causal relationship between an exposure variable and an outcome, given a set of assumptions.^21^ We employ it here to assess the evidence in support of causal effects of RNA expression of various genes on the odds of critical Covid-19.

We specified an *a priori* list of target genes that relate to the mechanism of action of many host-targeted drugs that have been proposed for the treatment of Covid-19 (Supp Table 1). Seven of these targets had a suitable locally-acting eQTL in GTEx(v7). Of these, *IFNAR2* remained significant after Bonferroni correcting for multiple testing for 7 tests (beta −1.49, standard error 0.52, p-value 0.0043), with equivocal evidence of heterogeneity (HEIDI^22^ p-value = 0.0150; 0.05/7 < p-value < 0.05; 6 SNPs). This Mendelian randomisation result successfully replicated in the results of COVID19-hg (ANA_B2_V2; hospitalized covid vs. population; UK Biobank excluded): beta −1.37, standard error 0.51, p-value 0.0066 (1 test).

We then performed transcriptome-wide Mendelian randomisation to quantify support for *unselected* genes as potential therapeutic targets. Instruments were available for 4,614 unique Ensembl gene IDs. No genes were statistically significant after correcting for multiple comparisons in this analysis (4,614 tests). After conservative filtering for heterogeneity (HEIDI p-value > 0.05), the smallest Mendelian randomisation p-value was 0.00049 for a variant at chr19:10466123 affecting expression of *TYK2*. 9 other genes with nominally significant Mendelian randomisation pvalues (p<0.005) were then tested for independent external evidence (Supplementary Information). We found that *TYK2* had a significant independent Mendelian randomisation p = 0.0022 in this second set (Bonferroni-corrected significance threshold = 0.006). In both cases it should be noted that this is not a complete replication, because the same GTEx expression data was used for both analyses.

### TWAS

We performed TWAS^23,24^ to link GWAS results to tissue-specific gene expression data by inferring gene expression from known genetic variants that are associated with transcript abundance (expression quantitative trait loci, eQTL). For this analysis we used GTExv8 data for two disease-relevant tissues chosen *a priori*: whole blood and lung(Figure 3), and performed a combined meta-TWAS analysis^25^ to detect predicted expression differences across both tissues.

**Figure 1:**
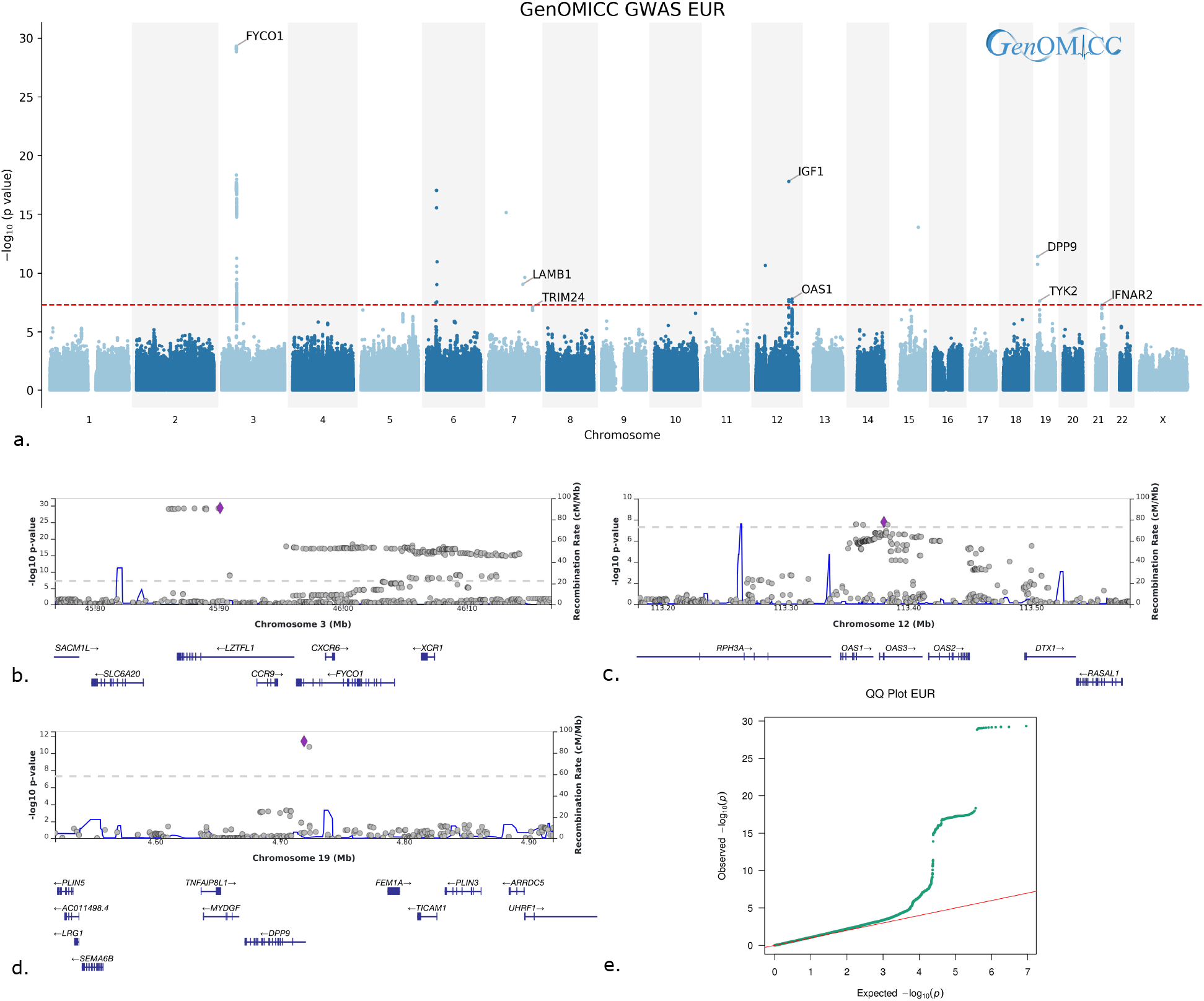
Summary of GWAS results for EUR ancestry group in GenOMICC. *a*. Manhattan plot showing SNP-level p-values for genome-wide significant associations in largest ancestry group, EUR (red horizontal line shows genome-wide significance at −log_10_ (5×10^−8^)) *b-d*. Locuszoom^19^ plots showing genomic regions around protein-coding genes. *e*. Quantile-quantile (QQ) plot.

**Figure 2:**
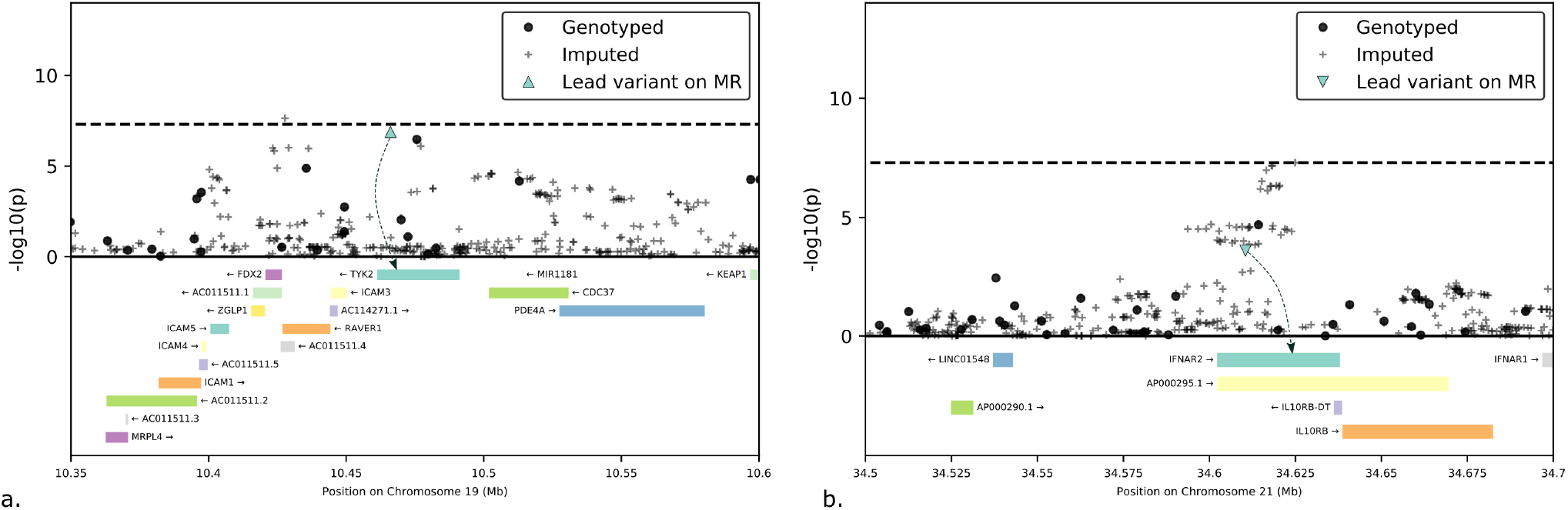
Detail of the TYK2 and IFNAR2 loci showing location of instrumental variables used for Mendelian randomisation. x-axes: genomic position with gene locations marked; y-axes: GenOMICC GWAS −log10(p-values); dashed line: genome-wide significance at −log_10_ (5×10^−8^); blue triangles: variants used as the instrumental variables for MR, for TYK2 and IFNAR2, respectively. A detailed description of the method of instrument selection is provided in Materials and Methods.

**Figure 3:**
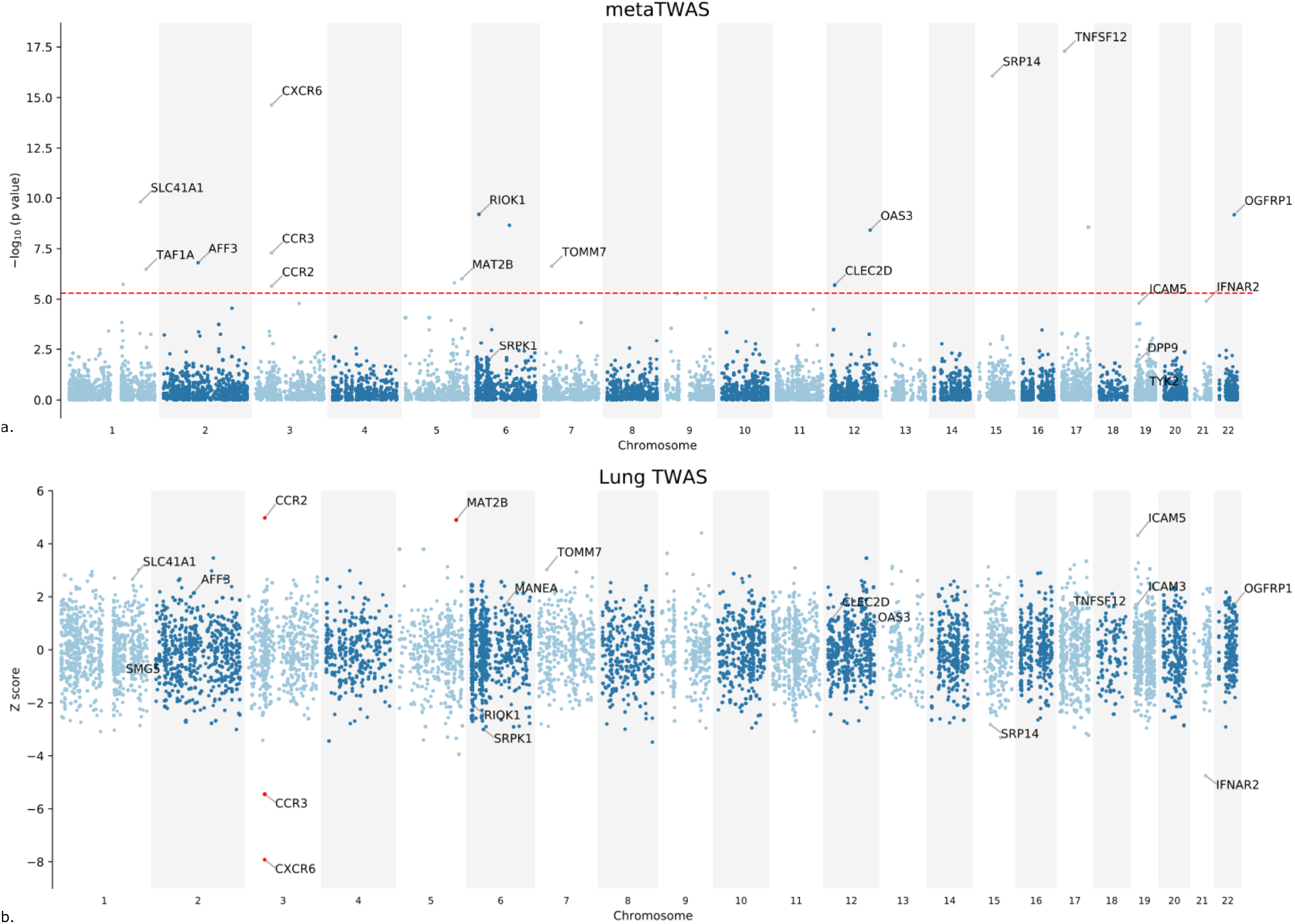
Summary of TWAS results. *a*. Gene-level Manhattan plot showing results from meta-TWAS analysis across tissues. *b*. z-scores showing direction of effect for genotype-inferred expression of transcripts encoding protein-coding genes in lung tissue (GTEXv8). Red highlighting indicates genome-wide significance at p < 5 × 10^−6^.

We discovered 18 genes with genome-wide significant differences in predicted expression compared to controls (Supplementary Information). This included 4 genes with differential predicted expression in lung tissue (Figure 3; 3 on chr3: *CCR2, CCR3* and *CXCR6*, and 1 on chr5: *MTA2B*).

#### Genetic correlations, tissue, and cell-type associations

We tested for genetic correlations with other traits, that is the degree to which the underlying genetic components are shared with severe Covid-19. Using the high-definition likelihood (HDL) method,^26^ we identified significant negative genetic correlations with educational attainment and intelligence. Significant positive genetic correlations were detected for a number of adiposity phenotypes including body mass index and leg fat, as well as pulse rate, neck/shoulder pain, thyroxine, and lansoprazole medications.

Consistent with GWAS results from other infectious and inflammatory diseases,^27^ there was a significant enrichment of strongly-associated variants in promoters and enhancers, particularly those identified by the EXaC study as under strong evolutionary selection (Supplementary Information).^28^ The strongest tissue type enrichment was in spleen, followed by pancreas (Supplementary Information).

## Discussion

We have discovered and replicated significant genetic associations with life-threatening Covid-19 (Figure 1). Our focus on critical illness increases the probability that some of these associations relate to the later, immune-mediated disease associated with respiratory failure requiring invasive mechanical ventilation.^2^ Importantly, the GWAS approach is unbiased and genome-wide, enabling the discovery of completely new pathophysiological mechanisms. Because genetic variation can be used to draw a causal inference, genetic evidence in support of a therapeutic target substantially improves the probability of successful drug development.^29^ In particular, Mendelian randomisation occupies a uinque position in the hierarchy of clinical evidence.^30^

Patients admitted to intensive care units in the UK during the first wave of Covid-19 were, on average, younger and less burdened by comorbid illness than the hospitalised population.^15^ Compared to other countries, UK ICU admission tends to occur at a higher level of illness severity,^31^ reflected in the high rate of invasive mechanical ventilation use in our cohort (73%; Table 1). Therefore, the population studied here are defined by their propensity to critical respiratory failure due to Covid-19. GenOMICC recruited in 208 intensive care units (covering more than 95% of UK ICU capacity), ensuring that a broad spread across the genetic ancestry of UK patients was included (Supplementary Information).

For external replication, the nearest comparison is the hospitalised vs population analysis in the Covid-19 Host Genetics initiative, which has been generously shared with the international community. Likewise, full summary statistics from GenOMICC are openly available in order to advance the rate of discovery.

Despite the differences in case definitions, novel associations from our study of critical illness replicate robustly in the hospitalised case study(Table 3). Separately, the Mendelian randomisation results implying a causal role for *IFNAR2* and *TYK2* are also statistically significant in this cohort. Our findings reveal that critical illness in Covid-19 is related to at two biological mechanisms: innate antiviral defences, which are known to be important early in disease (*IFNAR2* and *OAS* genes), and host-driven inflammatory lung injury, which is a key mechanism of late, life-threatening Covid-19 (*DPP9, TYK2* and *CCR2*).^2^

Interferons are canonical host antiviral signalling mediators, and stimulate release of many essential components of the early host response to viral infection.^32^ Consistent with a beneficial role for type I interferons, increased expression of the interferon receptor subunit *IFNAR2* reduced the odds of severe Covid-19 with Mendelian randomisation discovery p = 0.0043 (7 tests); replication p = 0.0066 (1 test). Within the assumptions of Mendelian randomisation, this represents evidence for a protective role for IFNAR2 in Covid-19. We deemed this gene to be therapeutically-informative *a priori* because it is a target for exogenous interferon treatment. with fatal sequelae from live-attenuated measles virus in humans^33,34^ and with influenza in mice.^35^

The variant rs10735079 (chr12, p = 1.65 × 10^−8^) lies in the oligoadenylate synthetase (OAS) gene cluster (*OAS1, OAS2* and *OAS3*; Figure 1). TWAS also (Figure 3) detects significant associations in these genes(Figure 3). OAS genes are inducible by type I interferons. These genes encode enzymes which activate an effector enzyme, RNAse L, which degrades double-stranded RNA,^36^ a replication intermediate of coronaviruses.^37^ *OAS1* variants were implicated in susceptibility to SARS-CoV in candidate gene association studies in Vietnam^38^ and China.^39^

The association in 19p13.3 (rs2109069, p = 3.98 × 10^12^) is an intronic variant in the gene encoding dipeptidyl peptidase 9 (*DPP9*). Variants in this locus are associated with idiopathic pulmonary fibrosis^40^ and interstitial lung disease.^41^ *DPP9* encodes a serine protease with diverse intracellular functions, including cleavage of the key antiviral signalling mediator CXCL10,^42^ and plays a key role in antigen presentation.^43^

Since opportunities for therapeutic intervention, particularly experimental therapy, are more abundant in later, more severe disease, it is important that our results also reveal genes that may act to drive inflammatory organ injury. *TYK2* is one of 4 gene products listed in the druggable genome Targets Central Resource Database^44^ as a target for baricitinib, one of the nine candidate drugs we used in the creation of our *a priori* target list (Supplementary Table 1). However, since we did not *a priori* include *TYK2* on the final set of genes for focused Mendelian randomisation, we use a significance threshold corrected for the full set of comparisons: discovery p = 0.00049 (4614 tests); replication p = 0.0022 (9 tests).

We replicate the finding of Ellinghaus *et al*. at 3p21.31.^11^ The extremely small p-value at this locus (p=4.77× 10^−30^) may reflect the strength of the signal, the large size of our study, our focus on extreme severity. The 3p21.31 locus is populated by a number of genes with mechanisms of action that could plausibly explain an association. Our systematic review and meta-analysis of experimental data on betacoronavirus infection from other sources provides moderate biological support for *FYCO1*, although the additional information comes mostly from *in vitro* model systems.^45^

TWAS results show that variants in this region confer genome-wide significant differences in predicted expression of *CXCR6, CCR2* and *CCR3* (Supplementary Information). Association with critical illness for genotype-inferred *CCR2* (CC-chemokine receptor 2) expression is particularly strong in lung tissue(Figure 3). CCR2 promotes monocyte/macrophage chemotaxis towards sites of inflammation, and there is increased expression of the canonical ligand for CCR2, monocyte chemoattractant protein (MCP-1), in bronchoalveolar lavage fluid from the lungs of Covid-19 patients during mechanical ventilation.^46^ Circulating MCP-1 concentrations are associated with more severe disease.^47^ Anti-CCR2 monoclonal antibody therapy in treatment of rheumatoid arthritis is safe.^48^

The *ABO* locus was also previously associated with Covid-19,^11^ but was not significant in our study (smallest *p*=1.30 × 10^−3^, chr9:136115876; Supplementary Information). This does not rule out the possibility of a true association, but other possible explanations include differences between case and control populations in each study.

Analysis of shared heritability highlights adiposity and educational attainment: genetically increased body mass index (BMI) and genetically decreased educational attainment are both associated with higher risk of severe Covid. This does not imply a causal relationship, as a number of biases may be at play, but may reflect the fact that increased BMI and lower socio-economic status are strong risk factors for Covid-19,^49,50^ or the fact that UK Biobank participants were disproportionately drawn from a high socio-economic status group.^51^

There is an urgent need to deepen these findings through further studies of this type, with harmonised integration across multiple studies. We continue to recruit to the GenOMICC study, in the expectation that additional associations exist and can be detected with larger numbers of cases. Future studies using whole genome sequencing will add spatial resolution on the genome and better detection rare variants. Effect sizes are likely to be higher in GenOMICC because the cohort is strongly enriched for immediately life-threatening disease in patients who are either receiving invasive mechanical ventilation, or considered by the treating physicians to be at high risk of requiring mechanical support. With 2244 cases we have statistical power to detect strong effects, such as the highly-significant locus at 3p21.31, as well as moderate genome-wide significant findings with external replication at *DPP9, OAS* and *IFNAR2*.

Because of the urgency of completing and reporting this work, we have drawn controls from population genetic studies who were genotyped using different technology from the cases. We mitigated the consequent risk of false-positive associations driven by genotyping errors by genotyping the majority of our subjects using two different methods, and by verifying significant associations using two separate control groups (100,000 genomes and Generation Scotland). The success of these mitigations is demonstrated by robust replication of our top hits in external studies.

We have discovered new and highly plausible genetic associations with critical illness in Covid-19. Some of these associations lead directly to potential therapeutic approaches to augment interferon signalling, antagonise monocyte activation and infiltration into the lungs, or specifically target harmful inflammatory pathways. While this adds substantially to the biological rationale underpinning specific therapeutic approaches, each treatment must be tested in large-scale clinical trials before entering clinical practice.

## Materials and methods

### Recruitment

2,636 patients recruited to the GenOMICC study (genomicc.org) had confirmed Covid-19 according to local clinical testing and were deemed, in the view of the treating clinician, to require continuous cardiorespiratory monitoring. In UK practice this kind of monitoring is undertaken in high-dependency or intensive care units. An additional 134 patients were recruited through ISARIC 4C (isaric4c.net) - these individuals had confirmed Covid-19 according to local clinical testing and were deemed to require hospital admission. Both studies were approved by the appropriate research ethics committees (Scotland 15/SS/0110, England, Wales and Northern Ireland: 19/WM/0247). Current and previous versions of the study protocol are available at genomicc.org/protocol.

### Genotyping

DNA was extracted from whole blood using Nucleon Kit (Cytiva) with the BACC3 protocol. DNA samples were re-suspended in 1 ml TE buffer pH 7.5 (10mM Tris-Cl pH 7.5, 1mM EDTA pH 8.0). The yield of the DNA was measured using Qubit and normalised to 50ng/µl before genotyping.

Genotyping was performed using the Illumina Global Screening Array v3.0 + multi-disease beadchips (GSAMD-24v3-0-EA) and Infinium chemistry. In summary this consists of three steps: (1) whole genome amplification, (2) fragmentation followed by hybridisation, and (3) single-base extension and staining. For each of the samples, 4 µl of DNA normalised to 50ng/µl was used. Each sample was interrogated on the arrays against 730,059 SNPs. The Arrays were imaged on an Illumina iScan platform and genotypes were called automatically using GenomeStudio Analysis software v2.0.3, GSAMD-24v3-0-EA_20034606_A1.bpm manifest and cluster file provided by manufacturer.

In 1667 cases, genotypes and imputed variants were confirmed with Illumina NovaSeq 6000 whole genome sequencing. Samples were aligned to the human reference genome hg38 and variant called to GVCF stage on the DRAGEN pipeline (software v01.011.269.3.2.22, hardware v01.011.269) at Genomics England. Variants were genotyped with the GATK GenotypeGVCFs tool v4.1.8.1,^52^ filtered to minimum depth 8X (95% sensitivity for heterozygous variant detection,^53^) merged and annotated with allele frequency with bcftools v1.10.2.

#### Quality control

Genotype calls were carefully examined within GenomeStudio using manufacturer and published^54^ recommendations, after excluding samples with low initial call rate (<90%) and reclustering the data thereafter. Briefly, X and Y markers calls were all visually inspected and curated if necessary, as were those for autosomal markers with minor allele frequency > 1% displaying low Gentrain score, cluster separation, and excess or deficit of heterozygous calls. Genotype-based sex determination was performed in GenomeStudio and samples excluded if not matching records expectation. Five individuals with XXY genotypes were also detected and excluded for downstream GWAS analyses. Genotypes were exported, in genome reference consortium human build 37 (GRCHb37) and Illumina “source” strand orientation, using the GenotypeStudio `plink-input-report-plugin-v2-1-4`. A series of filtering steps was then applied using PLINK 1.9 leaving 2799 individuals and 479095 variants for further analyses (exclusion of samples with call rate < 95%, selection of variants with call rate > 99% and minor allele frequency (MAF) > 1% and final samples selection using a call rate > 97%).

#### Kinship

Kinship and ancestry inference were calculated following UK Biobank^51^ and 1M veteran program.^55^ First King2.1^56^ was used to find duplicated individuals which have been recruited by two different routes. The analysis flagged 56 duplicated pairs, from which one was removed according to genotyping quality (GenomeStudio p50GC score or/and individual call rate). This leaves a set of 2734 unique individuals.

Regions of high linkage disquilibrium (LD) defined in the UK Biobank^51^ were excluded from the analysis, as well as SNPs with MAF<1% or missingness >1%. King 2.1 was used to construct a relationship matrix up to 3rd degree using the King command `--kinship --degree 3` and then the function `largest_independent_vertex_set()` from the `igraph` tool[[http://igraph.sf.net]] was used to create a first set of unrelated individuals. Principal component analysis (PCA) was conducted with gcta 1.9^57^ in the set of unrelated individuals with pruned SNPs using a window of 1000 markers, a step size of 80 markers and an r^2^ threshold of 0.1. SNPs with large weights in PC1, PC2 or PC3 were removed, keeping at least 2/3 of the number of pruned SNPs to keep as an input of the next round of King 2.1. The second round of King 2.1 was run using the SNPs with low weights in PC1, PC2 and PC3 to avoid overestimating kinship in non-european individuals. After this round 2718 individuals were considered unrelated up to 3rd degree.

#### Genetic ancestry

Unrelated individuals from the 1000 Genome Project dataset were calculated using the same procedure described above, and both datasets were merged using the common SNPs. The merged genotyped data was pruned with `plink` using a window of 1000 markers a step size of 50 and a r^2^ of 0.05, leaving 92K markers that were used to calculate the 20 first principal components with gcta 1.9. Ancestry for genomicc individuals was inferred using ADMIXTURE^58^ populations defined in 1000 genomes. When one individual had a probability > 80% of pertaining to one ancestry, then the individual was assigned to this ancestry, otherwise the individual was assigned to admix ancestry as in the 1M veteran cohort.^55^ According to this criterion there are 1818 individuals from European ancestry, 190 from African ancestry, 158 from East Asian ancestry, 254 from South Asian ancestry, and 301 individuals with admixed ancestry (2 or more).

### Imputation

Genotype files were converted to plus strand and SNPs with Hardy-Weinberg Equilibrium (HWE) p-value<10^−6^ were removed. Imputation was calculated using the TOPMed reference panel.^18^ and results were given in grch38 human reference genome and plus strand. The imputed dataset was filtered for monogenic and low imputation quality score (r^2^<0.4) using BCFtools 1.9. To perform GWAS, files in VCF format were further filtered for r^2^>0.9 and converted to BGEN format using QCtools 1.3.^59^

UK Biobank imputed variants with imputation score >0.9 and overlapping our set of variants (n=5,981,137) were extracted and merged with GenOMICC data into a single BGEN file containing cases and controls using QCtools 1.3.

### GWAS

Individuals with a positive Covid-19 test or suspected Covid-19 when they were admitted in the hospital were included in the GWAS as cases. Related individuals to degree 3 were removed. 13 individuals with American ancestry were removed as the sample size provided insufficient power to perform a reliable GWAS for this group. The final dataset includes 2244 individuals: 1676 individuals from European ancestry, 149 individuals from East Asian ancestry, 237 individuals from South Asian ancestry and 182 individuals from African ancestry (Table 1). If age or deprivation status were missing for some individuals, the value was set to the mean of their ancestry. GWAS were performed separately for each ancestry group.

Tests for association between case-control status and allele dosage at individuals SNPs were performed by fitting logistic regression models using PLINK.^60^ Independent analyses were performed for each ethnic group. All models included sex, age, mean centered age squared, deprivation score decile of residential postcode, and the first 10 genomic principal components as covariates.

Genomic principal components were computed on the combined sample of all UK Biobank and GenOMICC participants. Specifically, 456,750 genetic variants were identified which were shared between the variants contained in the called genotypes in the GenOMICC dataset and imputed UK Biobank genotypes, which had an information score above 0.95 and a minor allele frequency above 1%. After merging genotypes at these variants, variants were removed which had a minor allele frequency below 2.5%, a missingness rate above 1.5%, showed departure from Hardy-Weinberg equilibrium with a p value below 10^−50^, or which were within previously identified regions of high linkage disequilibrium within UK Biobank. After LD-pruning of the remaining variants to a maximum r^2^ of 0.01 based on a 1000 variant window moving in 50 variants steps, using the PLINK indep-pairwise command and yielding 13,782 SNPs, the leading 20 genomic principal components were computed using FlashPCA2.^61^

GWAS results were filtered for MAF>0.01, variant genotyping rate > 0.99 and HWE p-value > 10^−50^ for each ethnicity. An extra filter was added to avoid bias for using a different genotyping method and imputation panel between controls and cases. This could not be controlled for using regression because all cases and all controls were genotyped using different methods. MAF for each ancestry were compared between UK Biobank and gnomAD hg38 downloaded in August 2020.^62^ SNPs were were removed from the GWAS results specifically for each ethnicity following these two rules: (a) In SNPs with MAF > 10% in gnomAD, an absolute difference of 5% between gnomAD and UK biobank controls MAF (b) In SNPs with MAF <10% in gnomAD, a difference > 25% gnomAD MAF, between UK Biobank controls and gnomAD. To calculate differences between UK Biobank European individuals and gnomAD allele frequencies, non Finnish-europeans gnomAD allele frequencues were used, as European UK Biobank controls are mainly non-Finnish.

#### Deprivation score

The UK Data Service provides measures of deprivation based on Census Data and generated per postcode. The latest version of the Deprivation Scores were published in 2017 and are based on the 2011 census. Since only partial postcodes were available for most samples we were unable to use these indices directly. However, we generated an approximation to the scores by calculating an average weighted by population count across the top-level postcode areas.

The initial input file was part of the aggregated census data identified by DOI:10.5257/census/aggregate-2011-2.

Specifically the postcode data were downloaded from:

http://s3-eu-west-1.amazonaws.com/statistics.digitalresources.jisc.ac.uk/dkan/files/Postcode_Counts_and_Deprivation_Ranks/postcodes.zip

Population count and deprivation score for each published postcode were extracted and weighted average score calculated for each top-level postcode. We further categorised each top-level postcode score into decile and quintile bins for more coarse-grained analyses.

#### Whole Genome Sequencing

Whole Genome Sequencing (WGS) gVCF files were obtained for the 1667 individuals for which we had whole genome sequence data. Variants overlapping the positions of the imputed variants were called using GATk and variants with depth<8 (the minimum depth for which 95% coverage can be expected) were filtered. Individual VCF files were joined in a multi-sample VCF file for comparison with imputed variants. 1613 of these 1667 were used in the final GWAS. Samples were filtered and variants annotaed using bcftools 1.9. VCF files obtained from imputation were processed in an identical manner. Alternative allele frequency was calculated with PLINK 2.0^63^ for both WGS and imputed data.

### Controls

#### UK Biobank

UK Biobank participants were were considered as potential controls if they were not identified by the UK Biobank as outliers based on either genotyping missingness rate or heterogeneity, and their sex inferred from the genotypes matched their self-reported sex. For these individuals, information on sex (UKBID 31), age, ancestry, and residential postcode deprivation score decile was computed. Specifically, age was computed as age on April 1st, 2020 based on the participant’s birth month (UKBID 34) and year (UKBID 52). The first part of the residential postcode of participants was computed based on the participant’s home location (UKBID 22702 and 22704) and mapped to a deprivation score decile as previously described for GenOMICC participants. Ancestry was inferred as previously described for GenOMICC participants.

After excluding participants who had received PCR tests for Covid-19, based on information downloaded from the UK Biobank in August 2020, five individuals with matching inferred ancestry were sampled for each GenOMICC participant as controls. After sampling each control, individuals related up to 3rd degree were removed from the pool of potential further controls.

#### Generation Scotland

Generation Scotland: Scottish Family Health Study (hereafter referred to as Generation Scotland) is a population-based cohort of 24 084 participants sampled from five regional centers across Scotland(www.generationscotland.org).^64^ A large subset of participants were genotyped using either Illumina HumanOmniExpressExome-8v1_A or v1-2, and 20 032 passed QC criteria previously described.^65,66^ Genotype imputation using the TOPMed reference panel was recently performed (freeze 5b) using Minimac4 v1.0 on the University of Michigan serverhttps://imputationserver.sph.umich.edu.^67^ Imputation data from unrelated (genomic sharing identical by descent estimated using PLINK1.9 < 5%) participants were used as control genotypes in a GWAS using GenOMICC cases of European ancestry, for quality check purpose of associated variants.

### Replication

GenOMICC EUR loci were defined by clumping function of PLINK 1.9 and clumping parameters r2 0.1 pval=5e-8 and pval2 0.01, and distance to the nearest gene was calculated using ENSEMBL grch37 gene annotation.

No GWAS has been reported of critical illness or mortality in Covid-19. As a surrogate, to provide some replication for our findings, replication analyses were performed using Host Genetics Initiative build 37, version 2 (July 2020) B2 (hospitalised Covid-19 vs population) v2 GWAS. Summary statistics were used from the full analysis, including all cohorts and GWAS without UK Biobank, to avoid sample overlap. Replication p-value was set to 0.05/n, where n is the number of loci significant in the discovery.

### Post-GWAS analyses

#### TWAS

We performed transcriptome-wide association using the MetaXcan framework^25^ and the GTExv8 eQTL MASHR-M models available for download (http://predictdb.org/). First GWAS results were harmonised, lifted over to hg38 and linked to 1000 Genomes reference panel using GWAS tools https://github.com/hakyimlab/summary-gwas-imputation/wiki/GWAS-Harmonization-And-Imputation. TWAS for whole blood and lung were calculated using GWAS summary statistics for the European population GWAS and S-PrediXcan. Resulting p-values were corrected using the Bonferroni correction to find significant gene associations.

#### Mendelian randomisation

Two-sample Summary data based Mendelian randomisation [PMID 27019110] was performed using the results of GenOMICC and the Genotype-Tissue expression project, GTEx v7 (chosen in preference to v8 because of the availability of pre-computed data for SMR/HEIDI),[PMID 29022597] with Generation Scotland [PMID 22786799; PMID 17014726] forming a linkage disequilibrium reference. GenOMICC results from those of European ancestry were used as the outcome; and GTEx (v7) whole blood expression results as the exposure. Data pertaining to GTEx v7 chosen were downloaded from the GTEx portal - https://gtexportal.org/ (accessed 20 Feb 2020, 05 Apr 2020, and 04 Jul 2020), and SMR/HEIDI from https://cnsgenomics.com/software/smr/ (accessed 03 Jul 2020). Analyses were conducted using Python 3.7.3 and SMR/HEIDI v1.03. An LD reference was created using data from the population-based Generation Scotland cohort (used with permission; described previously [PMID 28270201]): from a random set of 5,000 individuals, using Plink v1.9 (www.cog-genomics.org/plink/1.9/), a set of individuals with a genomic relatedness cutoff < 0.01 was extracted; 2,778 individuals remained in the final set. All data used for the SMR/HEIDI analyses were limited to autosomal biallelic SNPs: 4,264,462 variants remained in the final merged dataset.

Significant (as per GTEx v7; nominal p-value below nominal p-value threshold) local (distance to transcriptional start site < 1Mb) eQTL from GTEx v7 whole blood for protein coding genes (as per GENCODE v19) with a MAF > 0.01 (GTEx v7 and GenOMICC) were considered as potential instrumental variables. Per variant, we first selected the Ensembl gene ID to which it was most strongly associated (so as to ensure that each variant can only be considered as an instrument for the gene to which it is most strongly associated) followed by selecting the variant to which each Ensembl gene ID was most strongly associated. Instruments were available for 4,614 unique Ensembl gene IDs.

Results were assessed based upon a list of genes selected *a priori* as of interest (Supplementary Table 1), and together as a whole. Partial replication of Bonferroni-corrected significant results was attempted in the results of COVID19-Host Genetics Initiative - https://www.covid19hg.org/ - with UK Biobank excluded (accessed 21 Sep 2020). Hospitalized covid vs. population (ANA_B2_V2) was selected as the phenotype most similar to our own, and therefore the most appropriate for use as a replication cohort. This is not a complete replication - due to the repeated use of GTEX v7 Whole Blood results in both analyses - yet remains informative as to the strength of assoiation between the genetic variant and COVID19, with a consistent Mendelian Randomisation effect-size estimate.

#### Gene-level

Gene-level burden of significance in the EUR ancestry group result was calculated using MAGMA v1.08.^68^ SNPs were annotated to genes by mapping based on genomic location. SNPs were assigned to a gene if the SNPs location is within 5 kb up- or down-stream of the gene region (defined as the transcription start site to transcription stop site). The MAGMA SNP-wise mean method was applied which utilises the sum of squared SNP Z-statistics as the test statistic. The 1000 Genomes Project European reference panel was used to estimate LD between SNPs.

Auxiliary files were downloaded from https://ctg.cncr.nl/software/magma on 1st September 2020. Gene location files for protein-coding genes were obtained from NCBI (ftp.ncbi.nlm.nih.gov):

`gene/DATA/GENE_INFO/Mammalia/Homo_sapiens.gene_info.gz`

on 29/04/2015, and from:

`genomes/Homo_sapiens/ARCHIVE/ANNOTATION_RELEASE.105/mapview/seq_gene.md.gz`

on 25/05/2016.

The reference data files used to estimate LD are derived from Phase 3 of the 1000 Genomes Project.

Competitive gene set enrichment analysis was conducted in MAGMA using a regression model that accounts for gene-gene correlations, to reduce bias resulting from clustering of functionally similar genes on the genome.^68^ Gene sets were queried from the databases KEGG 2019, Reactome 2016, GO Biological Process 2018, Biocarta 2016 and WikiPathways 2019. The Benjamini-Hochberg procedure was used to control false discovery rate (<0.05).

#### Meta-analysis by information content (MAIC)

Multiple *in vitro* and *in vivo* studies have identified key host genes that either directly interact with SARS-CoV-2, or define the host response to SARS-CoV-2. We have previously reported a systematic review of these studies.^45^ In order to put the new associations from this GWAS into context, we performed a data-driven meta-analysis of gene-level results combined with pre-existing biological data using meta-analysis by information content (MAIC).^69^ Briefly, MAIC combines experimental results from diverse sources in the form of ranked or unranked gene lists. The algorithm assigns a weighting to each input gene list, derived from the degree of overlap with other input lists. Each gene is then assigned a score calculated from the weightings for each gene list on which it appears. This process is repeated iteratively until all scores converge on a stable value. In order to prevent a single type of experiment from unduly biasing the results, input gene lists are assigned to categories, and a rule applied that only one weighting from each category can contribute to the score for any given gene.

#### Tissue and functional genomic enrichment

We downloaded the mean gene expression data summarised from RNA sequencing by the GTEx project (https://gtexportal.org/). The GTEx v7 data contain gene expressions of 19,791 genes in 48 human tissues. Gene expression values were normalized to numbers of transcripts per million reads (TPM). To measure the expression specificity of each gene in each tissue, each gene expression specificity was defined as the proportion of its expression in each tissue among all the tissues, i.e., a value ranging between 0 and 1. SNPs within the 10% most specifically expressed genes in each tissue were annotated for subsequent testing of heritability enrichment. For functional genomic enrichment analysis, we considered the inbuilt primary functional annotations v2.2 provided in the `ldsc` software (https://alkesgroup.broadinstitute.org/LDSCORE/) to annotated the SNPs.

With the annotated SNPs, we used stratified LD score regression (S-LDSC)^70^ to test whether any human tissue or specific functional genomic feature is associated with sereve Covid-19. Our GWAS summary statistics were harmonized by the munge_sumstats.py procedure in `ldsc`. LD scores of HapMap3 SNPs (MHC region excluded) for gene annotations in each tissue were computed using a 1-cM window. The enrichment score was defined as the proportion of heritability captured by the annotated SNPs divided by the proportion of SNPs annotated.

#### Testing genetic correlations with other phenotypes

We applied both the LD score regression (LDSC)^71^ and high-definition likelihood (HDL)^26^ methods to evaluate the genetic correlations between Severe Covid-19 and 818 GWASed phenotypes stored on LD-Hub.^72^ GWAS summary statistics were harmonized by the munge_sumstats.py procedure in the ldsc software. In the HDL analysis, we estimated the SNP-based narrow-sense heritability for each phenotype, and for the 818 complex traits GWASs, those with SNPs less than 90% overlap with the HDL reference panel were removed.

#### Genome build

Results are presented using Genome Reference Consortium Human Build 37. Imputed genotypes and whole-genome sequence data were lifted over from Genome Reference Consortium Human Build 38 using Picard liftoverVCF mode from GATK 4.0 which is based on the UCSC liftover tool(chain file obtained from

ftp://ftp.ensembl.org/pub/assembly_mapping/homo_sapiens/GRCh38_to_GRCh37.chain.gz.^73^

## Data Availability

Summary data will be available from the study website, https://genomicc.org/data and will be contributed to the Covid-19 human genetics innitiative.

## Acknowledgements

We thank the patients and their loved ones who volunteered to contribute to this study at one of the most difficult times in their lives, and the research staff in every intensive care unit who recruited patients at personal risk during the most extreme conditions we have ever witnessed in UK hospitals.

GenOMICC was funded by Sepsis Research (the Fiona Elizabeth Agnew Trust), the Intensive Care Society, a Wellcome-Beit Prize award to J. K. Baillie (Wellcome Trust 103258/Z/13/A) and a BBSRC Institute Program Support Grant to the Roslin Institute (BBS/E/D/20002172). Whole-genome sequencing was done in partnership with Genomics England and was funded by UK Department of Health and Social Care, UKRI and LifeArc. ISARIC 4C is supported by grants from: the Medical Research Council [grant MC_PC_19059], the National Institute for Health Research (NIHR) [award CO-CIN-01] and by the NIHR Health Protection Research Unit (HPRU) in Emerging and Zoonotic Infections at University of Liverpool in partnership with Public Health England (PHE), in collaboration with Liverpool School of Tropical Medicine and the University of Oxford [award 200907], NIHR HPRU in Respiratory Infections at Imperial College London with PHE [award 200927], Wellcome Trust and Department for International Development [215091/Z/18/Z], and the Bill and Melinda Gates Foundation [OPP1209135], and Liverpool Experimental Cancer Medicine Centre (Grant Reference: C18616/A25153), NIHR Biomedical Research Centre at Imperial College London [IS-BRC-1215-20013], EU Platform foR European Preparedness Against (Re-) emerging Epidemics (PREPARE) [FP7 project 602525] and NIHR Clinical Research Network for providing infrastructure support for this research. PJMO is supported by a NIHR Senior Investigator Award [award 201385]. The views expressed are those of the authors and not necessarily those of the DHSC, DID, NIHR, MRC, Wellcome Trust or PHE. HM was supported by the NIHR BRC at University College London Hospitals. The Health Research Board of Ireland (Clinical Trial Network Award 2014-12) funds collection of samples in Ireland.

This research has been conducted using the UK Biobank Resource under project 788. Generation Scotland received core support from the Chief Scientist Office of the Scottish Government Health Directorates [CZD/16/6] and the Scottish Funding Council [HR03006] and is currently supported by the Wellcome Trust [216767/Z/19/Z]. Genotyping of the GS:SFHS samples was carried out by the Genetics Core Laboratory at the Edinburgh Clinical Research Facility, University of Edinburgh, Scotland and was funded by the Medical Research Council UK and the Wellcome Trust (Wellcome Trust Strategic Award STratifying Resilience and Depression Longitudinally (STRADL) Reference 104036/Z/14/Z). Genomics England and the 100,000 Genomes Project was funded by the National Institute for Health Research, the Wellcome Trust, the Medical Research Council, Cancer Research UK, the Department of Health and Social Care and NHS England. Mark Caulfield is an NIHR Senior Investigator. This work is part of the portfolio of translational research at the NIHR Biomedical Research Centre at Barts and Cambridge. Research performed at the Human Genetics Unit was funded by the MRC (MC_UU_00007/10, MC_UU_00007/15). LK was supported by an RCUK Innovation Fellowship from the National Productivity Investment Fund (MR/R026408/1). ADB acknowledges funding from the Wellcome Trust PhD training fellowship for clinicians (204979/Z/16/Z), the Edinburgh Clinical Academic Track (ECAT) programme. We acknowledge support from the MRC Human Genetics Unit programme grant, “Quantitative traits in health and disease” (U. MC_UU_00007/10). A. Tenesa acknowledges funding from the BBSRC through programme grants BBS/E/D/10002070 and BBS/E/D/30002275, MRC research grant MR/P015514/1, and HDR-UK award HDR-9004 and HDR-9003.

This study owes a great deal to the National Institute of Healthcare Research Clinical Research Network (NIHR CRN) and the Chief Scientist Office (Scotland), who facilitate recruitment into research studies in NHS hospitals, and to the global ISARIC and InFACT consortia. We thank Dr. Jie Zheng (University of Bristol) for sharing the harmonized GWAS summary statistics used in LD-Hub.

